# Regular cannabis use is associated with altered neural and behavioural responses during anticipation and feedback of monetary reward and loss

**DOI:** 10.64898/2026.04.23.26351366

**Authors:** Giada Lombardi, Grace Blest-Hopley, Martina Maria Tarantini, Aisling O’Neill, Robin Wilson, Owen O’Daly, Vincent Giampietro, Sagnik Bhattacharyya

## Abstract

Regular cannabis use has been associated with alterations in reward-related neural processes, yet findings remain inconsistent and the relationship between neural activity and behavioural performance is not fully understood. The present study aimed to characterise neural and behavioural correlates of reward processing in regular cannabis users (CU) compared with matched non-users (NU) using the Monetary Incentive Delay Task (MIDT).

Firstly, we assessed behavioural performance through reaction times, accuracy and monetary earnings to determine whether potential neural alterations were reflected in task performance. Secondly, focusing on reward-related brain regions, we examined group differences in BOLD functional MRI activity during anticipation and outcome phases separately for monetary win and loss conditions. Finally, we explored the association between behavioural performance and neural activation. Our findings indicate that regular cannabis use is associated with altered engagement of key nodes within the mesocorticolimbic circuit during both anticipatory and outcome phases of reward processing, accompanied by impaired behavioural performance. Particularly, compared with NU, CU showed (I) lower striatal activity during anticipation of monetary win and higher ventral striatum and frontal pole activity during anticipation of monetary loss; (II) greater VTA activation during outcome of successful monetary win and loss avoidance and lower frontal pole activity during outcome of unsuccessful loss avoidance; (III) impaired behavioural performance, reflected in lower monetary rewards and a trend towards slower reaction times and reduced accuracy; (IV) disrupted brain-behaviour coupling.

Results from this study may help inform future research on the neurobiological mechanisms underlying changes in reward function and the resultant behavioural consequences of cannabis use.

## Introduction

Cannabis is the third most widely consumed drug after alcohol and nicotine, with an estimated 244 million users globally [1]. Notably, approximately one in ten users develop cannabis use disorder and are at heightened risk for adverse psychosocial outcomes, including cognitive impairment, mental health problems and hazardous behaviour [2-6]. Given the increasing prevalence of cannabis use and its notable psychosocial effects, a comprehensive understanding of cannabis-related behavioural and neurobiological mechanisms becomes increasingly critical. Such insight is essential not only to clarify potential long-term consequences, but also to guide safer and more effective translational applications of cannabinoid-based therapies, that are becoming more prevalent [7].

Accumulating evidence indicates that regular cannabis use is associated with disrupted reward processing [8]. These alterations involve key components of the mesocorticolimbic circuit, including the ventral tegmental area (VTA), striatum and prefrontal regions, which play a central role in motivational and reward-related signalling [9]. While acute cannabis use is associated with higher dopaminergic neurotransmission within the ventral striatum, heavy cannabis users are characterised by a hypodopaminergic reward system [10]. In animal studies, repeated exposure to delta-9-tetrahydrocannabinol (THC), the main psychoactive ingredient of cannabis, leads to neuroadaptations within the reward circuit [11]. In humans, positron emission tomography (PET) studies have reported reduced striatal dopamine synthesis in chronic cannabis users, which has been linked to reduced reward sensitivity and motivation [12,13].

Neuroimaging studies have commonly relied on the Monetary Incentive Task (MIDT) to investigate reward processing in health and disease, as it allows to dissociate neural responses during anticipation of monetary win and loss from those during outcome feedback [14]. Since the original investigations of the MIDT in healthy individuals, results reported that anticipation of reward was associated with increased activation of the ventral striatum, a key region involved in encoding motivational salience and reward prediction [15-17]. In contrast, findings from studies applying the MIDT to cannabis users have been heterogeneous. While some investigations have shown blunted striatal responses during reward anticipation, others have found no significant differences compared with non-users [8,18]. Notably, the majority of studies reported results of anticipation or outcome of win and loss alone, limiting a comprehensive understanding of how these neural processes interact and differentiate within cannabis users. In addition, limited research has explored the relationship between neural activations and behavioural performance, leaving it unclear whether altered reward-related brain responses reflect disrupted motivational salience attribution.

On this basis, the present study aimed to provide a more comprehensive characterization of reward processing in regular cannabis users (CU) compared with age- and sex-matched non-users (NU). Towards this purpose, we used the MIDT to examine behavioural and neural responses during anticipation and outcome phases separately for win and loss conditions. Firstly, we assessed behavioural performance through reaction times, accuracy and monetary earnings to determine whether potential neural alterations were reflected in task performance. Secondly, using a region-of-interest (ROI) approach focusing on key reward-related brain regions, we examined group differences in neural activation during anticipation and outcome of monetary win and loss. Finally, we explored the association between behavioural performance and neural activation.

## Methods and materials

### Participants and MRI acquisition

Participants were recruited using local and targeted online advertising. Ethical approval was obtained from the King’s College London Research ethics committee (PNM RESC HR-15/16–2416). All participants gave written informed consent prior to the study and were reimbursed for their time and expenses. Full sociodemographic data and recent drug history of the participants have been previously described in detail [19]. From this cohort, twenty Cannabis Users (CU) and twenty age and sex matched Non-Users (NU) completed task-fMRI acquisition of the Monetary Incentive Delay Task and were included in the present study (see Table 1 for details of this sub-cohort). Inclusion criteria for the CU group required participants to have been consuming cannabis on at least four or more days per week, for the 2 years prior to taking part in the study. Furthermore, they were required to have initiated regular cannabis use before the age of 18, defined as using cannabis more than twice monthly and on at least ten lifetime occasions [20]. Inclusion criteria for the NU group required participants to have used cannabis fewer than ten times in their lifetime before taking part in the study [20]. Exclusion criteria for both groups included history of a neurological disorder, diagnosis of a mental illness, being a recipient of psychiatric services, family history of psychosis in a first-degree relative, educational attainment suggestive of an IQ of less than 70, or any contradiction to MRI acquisition. All participants underwent a screening interview, in which they were asked about personal and family history of suffering from or receiving help/treatment for mental disorders, and a urine drugs screening (for amphetamine, cocaine, opiates, THC, phencyclidine, benzodiazepines, barbiturates, methadone, propoxyphene) on the day of MRI scanning. Participants were asked to refrain from using cannabis or alcohol on the day of MRI scanning, from caffeine intake for four hours, and from tobacco use for two hours before the scan.

**Table 1.** Socio-demographic and cannabis use descriptives. CU: cannabis users; NU: non-users.

|  | CU | NU | Statistics |
| --- | --- | --- | --- |
| **Sample size (n)** | **20** | **20** |  |
| **Age (mean ± SD, years)** | **25.0 (±3.7)** | **24.2 (±4.2)** | **p=0.55** |
| **Sex (% males)** | **55** | **60** |  |
| **Education (mean ± SD, years)** | **15.2 (±3.5)** | **17.4 (±2.4)** | **p=0.023** |
| **Cannabis Use descriptive** |  |  |  |
| **Age of onset (mean)** | **14.8** |  |  |
| **Years of use (mean)** | **10.3** |  |  |
| **Cost per week (mean, GBP)** | **50.8** |  |  |
| **Total lifetime joints (mean, n)** | **4660.7** |  |  |

### MRI acquisition

All MRI images were acquired using a GE SIGNA HDx 3T MRI scanner system (GE healthcare Milwaukee, USA) at the Centre for Neuroimaging Sciences, King’s College London. T2*-weighted images were acquired axially in 39 slices (3 mm) with a 0.3-mm slice gap (matrix size 64 × 64 voxels, in-plane voxel size 3.75 × 3.75mm). A 30-ms echo time, 90° flip angle and compressed acquisition with a 2-s repetition time and 3 s silence were also used. A high-resolution gradient echo image was acquired for co-registration and to help map the fMRI data onto standard space with 43 × 3 mm slices with a 0.3-mm slice gap (matrix size 128 × 128 voxels, in –plane voxel size 1.875 × 1.875 mm). A 30-ms echo time, 90° flip angle and repetition time of 3 s were used.

### Monetary Incentive Delay Task (MIDT)

Participants completed two consecutive 8-minutes runs of the Monetary Incentive Delay Task (MIDT, see Figure 1). The task included four reward conditions, indicated by learned visual cues displayed for 250 ms): neutral (square cue; £0.00), win (circle cue with varying amount of lines; small reward: £0.20, large reward: £2.00) and loss (inverted triangle cue, -£2.00). Participants were then presented with an anticipatory period indicated by a fixation cross on the screen for 3700-4500 ms followed by a target cue (filled triangle) during which they were instructed to press a button as quickly as possible to win money (150-300 ms). Any response occurring less than 100□ms after target onset was regarded as an unsuccessful “false start”. At the end of each trial, participants were presented with a feedback screen indicating both the amount of money won/lost in that specific trial (top number) and the participant’s total amount of money (bottom number). Each run consisted of a total of 48 trials (12 trials x 4 conditions). All participants began with an initial amount of £10.00 and received payment according to the total monetary rewards accumulated across the two runs on each study day. Prior to scanning, participants underwent standardized task training.

**Figure 1.**
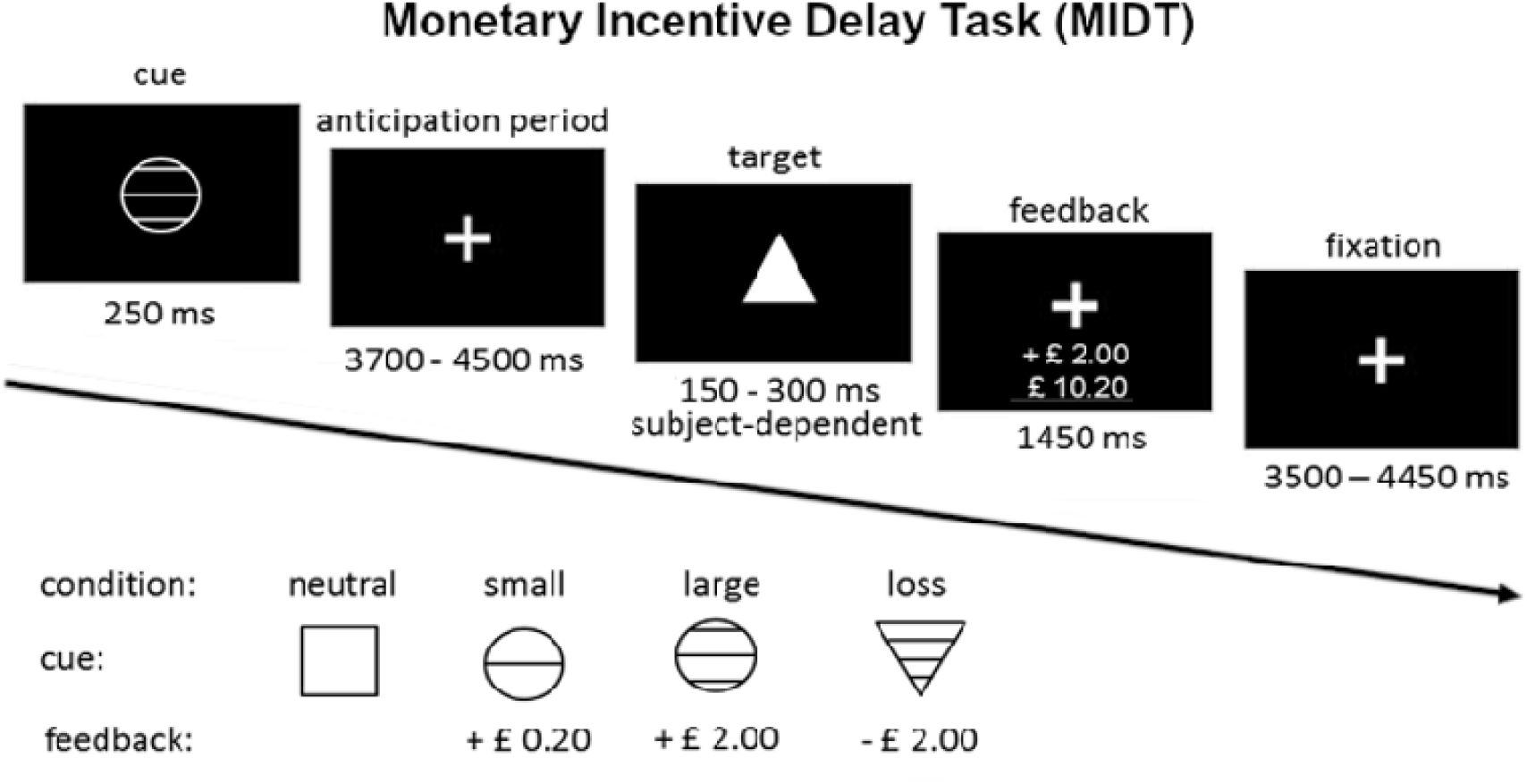
Monetary Incentive Delay Task (MIDT) paradigm.

### Performance analysis

Task performance was evaluated by calculating *mean monetary reward* earned during the task (£*), mean accuracy* (%), i.e. correct responses on target and the *mean reaction time (*RT, ms), i.e. the time occurring between target onset and participants’ response. Specifically, we computed both total scores of mean monetary reward and mean accuracy (all conditions collapsed together) and mean values of accuracy and RT for each condition (Win, Loss, Neutral). To test differences of accuracy and RT across the three conditions within the control group of NU we first computed a one-way ANOVA (see Supplementary Figure 1). Two independent two sample t-tests were used to compare the difference in mean monetary reward and total percentage of accurcy between CU and NU. Then, two-way factorial mixed design ANOVA was carried out to evaluate differences of accuracy and RT between groups (CU, NU) and across conditions (Win, Loss, Neutral).

### Task fMRI preprocessing and analysis

fMRI data were pre-processed and analysed with the SPM12 software running on MATLAB R2025b [21]. Preprocessing consisted of a standard pipeline including: (1) *realignment and unwarping* for motion and susceptibility distortion correction, (2) *slice timing* for the correction of slice-to-slice differences in time of BOLD signal sampling, (3) *normalization* into standard MNI152 space, (4) *functional smoothing* by using spatial convolution with a Gaussian kernel (FWHM = 8 mm). A general linear model (GLM) approach was employed to analyse task fMRI data, with regressors aligned with specific conditions of interest and convolved with the canonical hemodynamic response function. At the first level (single-subject level analysis), a model was designed and estimated for each participant to characterize neural responses during: 1) Anticipation of monetary win (*Win*), in which we collapsed together regressors associated with anticipation of winning a small or big reward, (2) Anticipation of monetary loss (*Loss*), (3) Feedback for successful win (*Win Hit*), (4) Feedback for successful loss avoidance (*Loss Hit)*, (5) Feedback for unsuccessful win (*Win Miss*), (6) Feedback for unsuccessful loss avoidance (*Loss Miss)*.

Each reward-related condition was contrasted with the corresponding neutral condition, which served as a baseline to isolate brain responses specifically related to reward anticipation and feedback. At the second level (group-level analysis), brain activation was examined using a region of interest (ROI) analysis (hypothesis-driven approach). ROIs were defined a priori based on key reward-related brain regions. Specifically, masks of dorsal striatum (encompassing caudate and putamen), ventral striatum and frontal pole were derived from the Harvard-Oxford atlas, and a mask of the ventral tegmental area (VTA) was obtained from the Harvard AAN atlas [22]. The BOLD signal (mean beta values) during the experimental conditions of interest was then extracted from each ROI for each participant. Group comparisons between CU and NU were then carried out using paired-samples t-tests, with a significance threshold set at p<0.05. We also conducted an exploratory whole-brain analysis in SPM (see Supplementary material).

### Correlation analysis

To assess whether increased or decreased brain activity during anticipation of win or loss was associated with faster or slower reaction times (RTs), we computed Pearson’s correlation coefficients between BOLD signal and RTs. Specifically, the BOLD signal during the anticipation conditions (*Win, Loss*) was extracted from ROIs used in the fMRI analysis and correlated with the corresponding condition-specific RTs. Differences between correlation coefficients of the two groups were assessed using Fisher’s r-to-z transformation for independent correlations.

## Results

### Behavioural performance

Results of the independent two sample t-tests revealed a significant difference between CU and NU of mean monetary reward won (p=0.04, Figure 2 A1) and a difference of total mean accuracy close to significance (p=0.06, Figure 2 A2).

**Figure 2.**
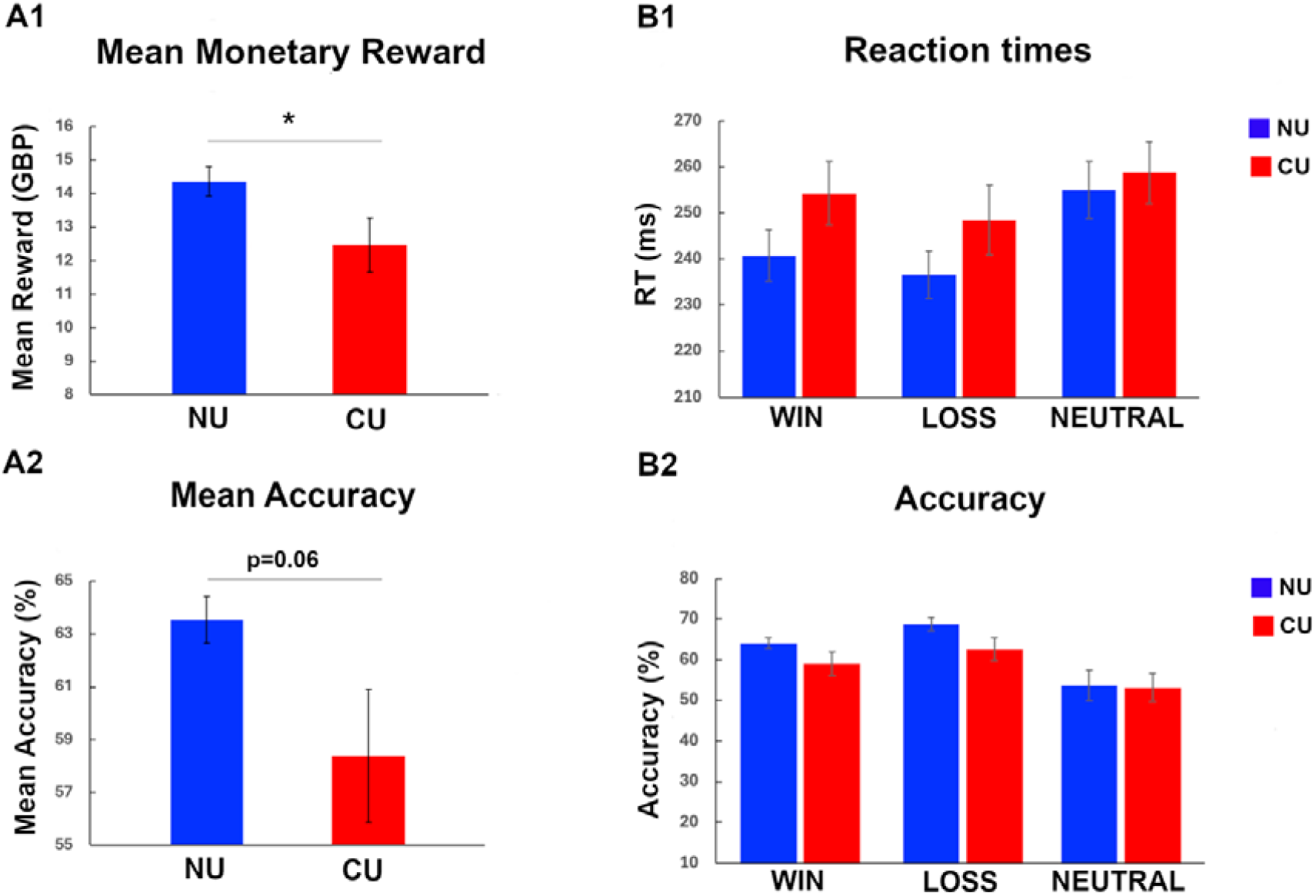
Results of mean monetary reward (A1) and total mean accuracy (A2) comparisons between CU and NU. Results of comparison across conditions and between groups for reaction times (B1) and accuracy (B2). Red bars represent cannabis users (CU) values. Blue bars represent non-cannabis users (NU) values. Vertical bars represent standard error of the mean (SEM). Horizontal bar represents statistical significance (*p<0.05).

For RT, results of the two-way factorial mixed design (Figure 2 B1) showed a significant main effect of Condition (F=21.41, p<0.001, ηp^2^=0.36). Specifically, Bonferroni-corrected pairwise comparisons revealed a significant difference in RT between *Win* and *Neutral* (p=0.001), *Loss* and *Neutral* (p<0.001), and *Win* and *Loss* conditions (p=0.004). Additionally, results revealed no significance for the main effect of Group (F= 1.25, p=0.27, ηp^2^=0.03) and an interaction between Group and Condition close to significance (F=2.75, p=0.071, ηp^2^=0.067).

For Accuracy, results of the two-way factorial mixed design (Figure 2 B2) showed a significant main effect of Condition (F=12.11, p<0.001, ηp^2^=024). Specifically, Bonferroni-corrected pairwise comparisons revealed a significant difference in RT between *Win* and *Neutral* (p=0.012) and between *Loss* and *Neutral* (p<0.001), but no significant difference beween *Win* and *Loss* conditions (p=0.184). Additionally, results revealed no significance for both the main effect of Group (F=2.89, p=0.097, ηp^2^=0.07) and the interaction between Group and Condition (F=0.9, p=0.386, ηp^2^=0.023).

### Task fMRI

Results of the exploratory whole brain analysis revealed an extended activation pattern for the control group of NU (see Supplementary material).

Figure 3 showed results of the main ROI-based analysis. During anticipation of monetary reward (*Win* > *Neutral*), NU exhibited robust activation in both the ventral and dorsal striatum. Group comparisons revealed that CU showed reduced activation in these regions, with significant differences observed in the ventral (p=0.008) and dorsal striatum (p=0.035). During anticipation of monetary loss (*Loss*>*Neutral*), NU again showed greater striatal engagement. Group comparisons indicated that CU exhibited lower BOLD signal in the ventral striatum, with the difference approaching significance (p=0.09).

**Figure 3.**
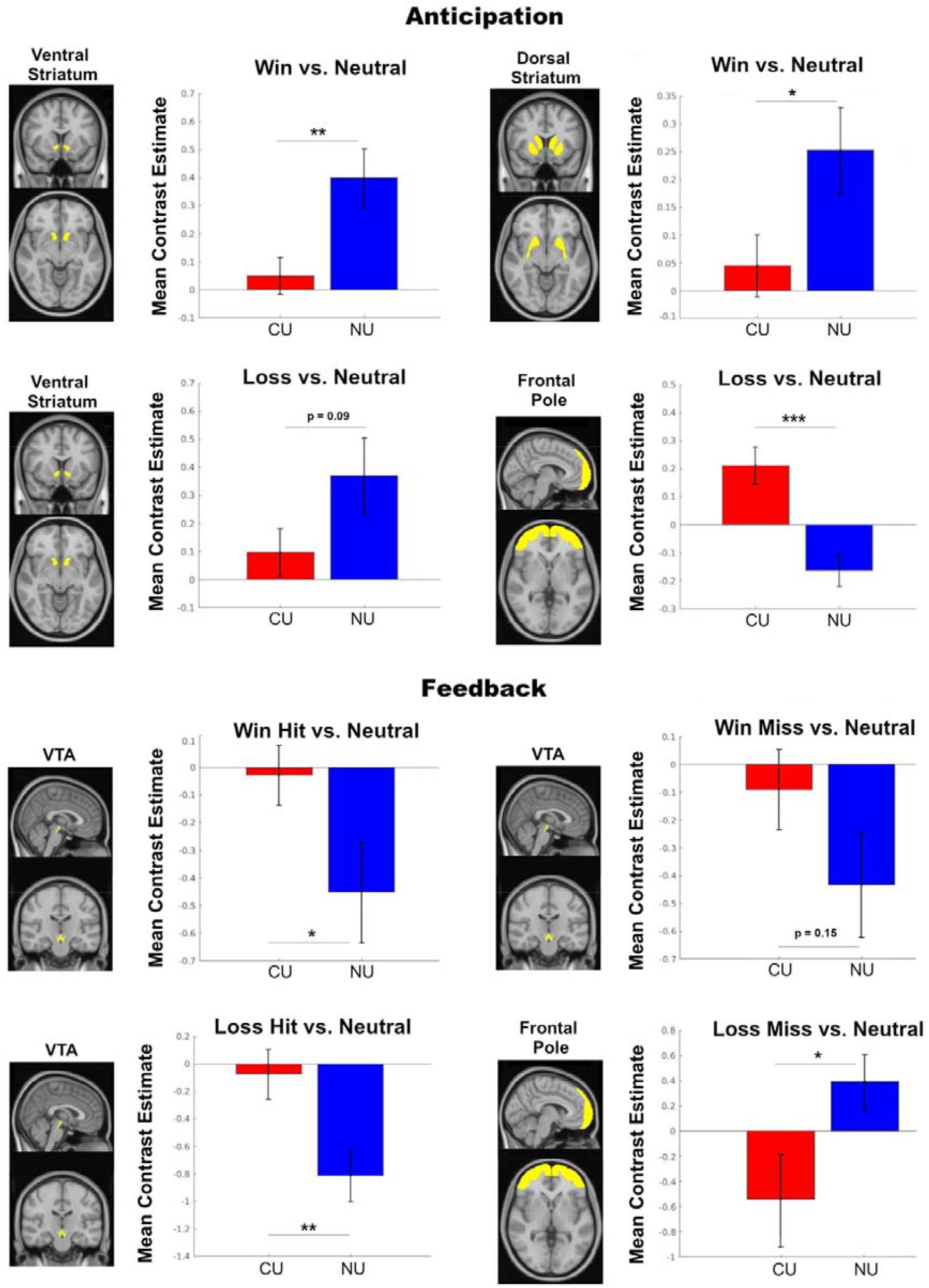
Results of the ROI analysis. Bar plots display the mean contrast estimates (β values) for each ROI for cannabis users (CU, red bars) compared to non-users (NU, blue bars). Vertical bars represent standard error of the mean (SEM). Horizontal bars indicate statistical significance (*p.0.05, **p.0.01, ***p.0.001).

Moreover, during *Loss*>*Neutral*, NU exhibited a negative BOLD response in the frontal pole. In contrast, CU showed a positive signal in the same region, and the difference between groups was statistically significant (p<0.001).

During the feedback phase, NU showed a pronounced negative BOLD response in the VTA. Group comparisons revealed differences of VTA activation between CU and NU for outcomes of successful win (*Win Hit>Neutral*, p=0.05) and loss avoidance (*Loss Hit>Neutral*, p=0.0065), and a trend of significance for outcome of unsuccessful win (*Win Miss>Neutral*, p=0.15). During outcome of unsuccessful loss avoidance (*Loss Miss*>*Neutral*), a significant group difference was observed in the frontal pole (p=0.035), with NU showing a positive BOLD response and CU exhibiting a robust negative BOLD response.

### Correlation analysis

For the NU group, Pearson’s correlation analyses revealed significant correlations between BOLD responses during the anticipation conditions and RTs. Specifically, we found a negative correlation between BOLD activity of the ventral striatum and RT of *Win* (Figure 4 A1, r=-0.62 , p=0.003) and *Loss* (Figure 4 A2, r=-0.57 , p=0.009) conditions and a negative correlation between BOLD activity of the dorsal striatum and RT of *Win* condition (Figure 4 A3, r=-0.6, p=0.005). In contrast, no significant correlations were observed for the CU group. Fisher’s r-to-z transformation analysis revealed significant differences between groups: (i) BOLD activity of ventral striatum – RT of *Win*: r_NU_=-0.62, r_CU_=0.09, p=0.01; (ii) BOLD activity of ventral striatum – RT of *Loss*: r_NU_=-0.57, r_CU_=0.02, p=0.05; (iii) BOLD activity of dorsal striatum – RT of *Win*: r_NU_=-0.6, r_CU_=0.31, p=0.003.

**Figure 4.**
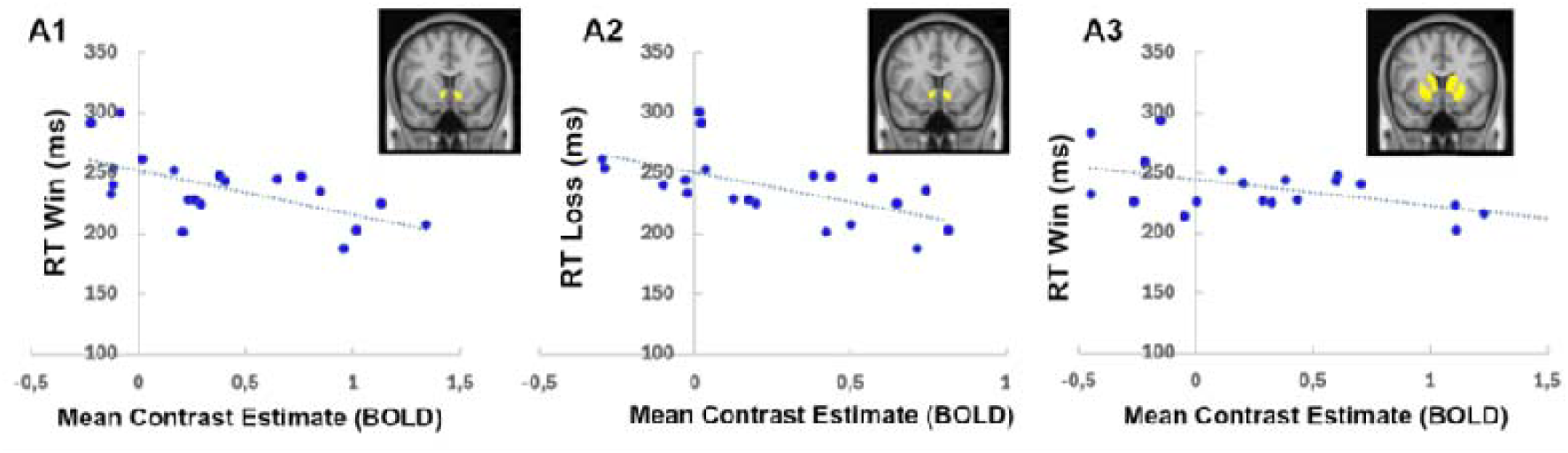
Significant negative correlations between BOLD activity of (A1) ventral striatum during Win anticipation, (A2) ventral striatum during Loss anticipation, (A3) dorsal striatum during Win anticipation and condition-specific reaction times (RT).

## Discussion

In the present study, we investigated brain and behavioural correlates of reward processing in regular cannabis users (CU) compared with age and sex matched non-users (NU) using the Monetary Incentive Delay Task (MIDT).

Behavioural results confirmed a robust effect of condition, with faster reaction times and higher accuracy in motivationally salient conditions (*Win* and *Loss*) compared with *Neutral* trials, indicating that task performance was effectively modulated by reward. Importantly, no significant main effect of group or interaction between group and condition was observed for either reaction times or accuracy. However, a trend towards a group × condition interaction was observed for reaction times, alongside a consistent pattern of slower responses and lower accuracy in CU compared with NU across conditions. Although these differences did not reach statistical significance, they may suggest a general impairment in task performance in CU.

One possible interpretation is that monetary reward may be less salient or motivating for regular CU. In line with incentive sensitization models of addiction [23], repeated exposure to drug-related rewards may bias motivational systems towards drug-related stimuli, resulting in reduced behavioural responsiveness to non-drug reward cues such as those used here. Alternatively, based on previous findings demonstrating that cannabis use is associated with slower reaction times [24-26], the observed pattern may reflect more general psychomotor slowing associated with chronic cannabis use. This slowing may partly reflect a compensatory strategy, as individuals under the influence of cannabis adopt a more cautios approach due to awareness of increased likelihood of errors. However, in the present study, accuracy remained lower in CU compared with NU, suggesting that such compensatory adjustments may be insufficient to fully mitigate performance deficits. Importantly, despite the absence of a significant group interaction, CU still showed modulation of behaviour across conditions, suggesting that motivational cues were processed, albeit less efficiently than in NU. Taken together, these results point to a subtle but consistent alteration in behavioural performance in CU, which is further supported by our neuroimaging findings.

Compared to NU, CU exhibited lower striatal activity (both ventral and dorsal striatum) during anticipation of monetary win, as well as lower ventral striatum activity and higher frontal pole activity during anticipation of monetary loss. The ventral striatum has been consistently implicated in reward anticipation and reward prediction error signalling [14, 27-28], primarily mediated by dopaminergic projections from the ventral tegmental area (VTA). This region represents a key hub for the acquisition and development of reward-based behaviours, including drug addiction and seeking [29-30]. Chronic cannabis use has been associated with blunted ventral striatal activity during anticipation of rewards [18], potentially reflecting a downregulation of CB1 receptors, which may disrupt dopaminergic neurotransmission within the reward circuit [10, 31-33]. Furthermore, a longitudinal study showed that greater cannabis use predicted attenuated ventral striatum responses during reward anticipation [34]. In addition to the ventral striatum, CU also showed reduced activation of the dorsal striatum during anticipation of monetary win compared with NU. The dorsal striatum is critically involved in translating motivational signals into goal-directed actions and in forming stimulus–response–reward associations that guide behaviour [10,35]. Dopaminergic projections to this region are thought to facilitate the integration of motivational value into action selection and reinforcement learning processes [27]. Cannabis use may disrupt this mechanism, resulting in reduced engagement of dorsal striatum during reward anticipation and potentially diminishing sensitivity to motivationally relevant cues. In addition, during the anticipation of monetary loss CU exhibited increased frontal pole activity, which may reflect enhanced cognitive control demand and evaluating processes when anticipating potentially negative outcomes. This activation pattern may represent a compensatory mechanism, whereby increased recruitment of control-related regions occurs in response to reduced efficiency of striatal activity. Similar compensatory engagement of frontal areas has been observed across substance use disorders and has been interpreted as reflecting increased cognitive effort to regulate behaviour in the context of altered reward system functioning [33,36].

Notably, while NU demonstrated a significant correlation between striatal activation during reward anticipation and reaction times, this relationship was not significant in CU. The absence of this brain–behaviour association further suggests a functional decoupling between neural reward signalling and behavioural output in CU, potentially reflecting impaired integration of motivational signals into motor preparation processes.

During the outcome feedback of successful win and loss avoidance, CU exhibited increased activity of the ventral tegmental area (VTA) compared with NU. VTA is a key component of the mesocorticolimbic dopaminergic system and plays a central role in encoding reward prediction errors, which signal the discrepancy between expected and actual outcomes [37]. Dopaminergic neurons in the VTA exhibit increased phasic activity in response to motivationally salient outcomes, including both rewarding and unexpected events, supporting reinforcement learning and facilitating future behavioural adaptation [38]. The increased VTA activity observed in CU during outcome feedback may reflect altered dopaminergic signalling dynamics associated with chronic cannabis exposure. Cannabinoids, primarily via CB1 receptor activation, modulate dopaminergic neurotransmission indirectly by acting on GABAergic and glutamatergic inputs to VTA dopaminergic neurons, resulting in their disinhibition and increased dopamine release [39]. The reduced anticipatory striatal activity observed in CU may result in weaker encoding of expected outcomes. Consequently, feedback signals may generate larger prediction errors, leading to enhanced dopaminergic responses and increased VTA activation during outcome evaluation. Interestingly, group differences were also observed in frontal pole activity during the outcome of unsuccessful loss avoidance. The frontal pole is involved in higher-order cognitive processes, including prospective evaluation, risk monitoring, integration of motivational and emotional information to guide behaviour, particularly in situations requiring evaluation of alternative courses of action, and adaptive decision-making under uncertainty [40-43]. The positive activation observed in NU may reflect efficient engagement of these processes, supporting adaptive behavioural adjustment following monetary loss. In contrast, the negative BOLD response in CU may indicate a reduced or maladaptive engagement of frontal mechanisms involved in processing negative feedback. This pattern could reflect diminished sensitivity to negative outcomes or an impaired ability to update behaviour based on adverse consequences. Future studies may investigate whether this underlies repetitive patterns of cannabis use despite harmful consequences. Notably, this finding complements the increased frontal pole activation observed during loss anticipation in CU. Such a dissociation is consistent with models of addiction suggesting that reduced anticipatory engagement of reward-related regions may be accompanied by heightened neural responses during reward receipt, reflecting altered reward learning and valuation processes [33].

It is important to note that CU were required to have initiated regular use before the age of 18, a developmental period characterised by heightened vulnerability to exogenous neurobiological insults due to ongoing brain maturation and endocannabinoid system development. Therefore, the present findings may partly reflect the effects of cannabis exposure during a particularly sensitive neurodevelopmental window.

The present work has several strengths. First, we employed a well-validated paradigm (MIDT) that allows the dissociation of neural responses during anticipation and outcome phases, enabling a deep investigation of win and loss processing in CU. Second, we separately examined win and loss conditions and assessed both behavioural performance and neural activation, providing a comprehensive characterisation of reward-related mechanisms in regular CU. Third, the inclusion of brain–behaviour correlations allowed us to link neural alterations with task performance. However, some limitations should also be considered. First, the cross-sectional design precludes causal inference regarding the directionality of the observed association between cannabis use and alterations of reward processing. Longitudinal studies will be necessary to confirm whether these neural and behavioural differences represent a consequence of chronic cannabis exposure. Second, the sample size was relatively small, limiting statistical power and generalisability of findings. Replication in larger cohorts is warranted.

In summary, the present study provides evidence that regular cannabis use is associated with altered behavioural and neural responses during reward processing. Due to the cross-sectional nature of the study, it remains unclear whether these results reflect neuroadaptations caused by repeated cannabis exposure or by differences between the two groups that may have predisposed CU to cannabis use. Clarifying this causal direction will be crucial to establish whether the observed differences are consequence or cause of cannabis use, thereby informing prevention strategies and clinical interventions targeting motivational and reward-related dysfunction.

## Data Availability

All data produced in the present study are available upon reasonable request to the authors

## Author contribution

GL performed data analysis and wrote the first draft of the manuscript; GBH contributed to data collection and analysis; MMT contributed to data analysis and to writing the first draft of the manuscript; AO’N. and RW contributed to data collection. OO’D and VG contributed to data interpretation; SB conceptualized the study, supervised the development of the study and contributed to data interpretation. All co-authors read the manuscript and approved its final version.

