## Supplementary material for "Regular cannabis use is associated with altered neural and behavioural responses during anticipation and feedback of monetary reward and loss"

For RT, results of the one-way ANOVA for the NU group showed a significant main effect of Condition (F=24.7, p<0.001, ηp²=0.36, see Supplementary Figure 1). Bonferroni-corrected pairwise comparisons revealed a significant difference in RT between *Win* and *Neutral* (p<0.001), *Loss* and *Neutral* (p<0.001), and *Win* and *Loss* conditions (p=0.007).

For accuracy, results of the one-way ANOVA for the NU group showed a significant main effect of Condition (F=8.15, p=0.001, ηp²=0.3, see Supplementary Figure 1). Bonferroni-corrected pairwise comparisons revealed a significant difference in accuracy between *Loss* and *Neutral* (p=0.006), but no significant difference between *Win* and *Neutral* (p=0.065) and between *Win* and *Loss* (p=0.82).

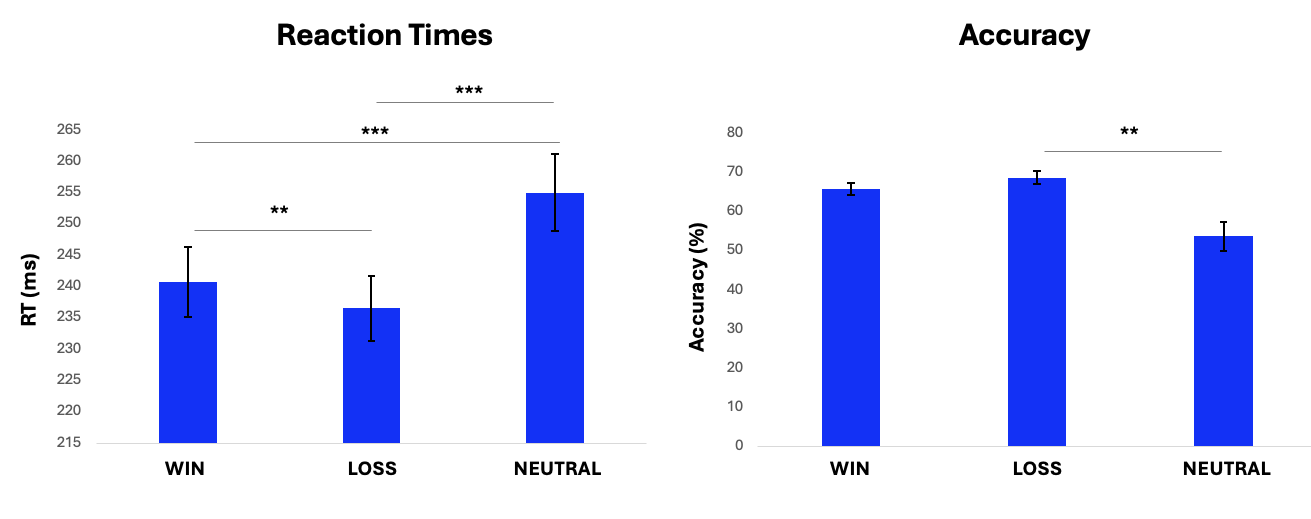

**Supplementary Figure 1. Results of reaction times and total mean accuracy comparison across conditions for the NU group. Vertical bars represent standard error of the mean (SEM). Horizontal bars represent statistical significance (**p<0.01; ***p<0.001).**

To complement the ROI-based analysis, we carried out an exploratory whole-brain analysis in SPM to examine task-related activation patterns in the control group NU (Supplementary Figure 2) and group differences (Supplementary table 1) in conditions of interest. Results were considered significant if they survived a family-wise error (FWE) correction of p<0.05 on the basis of cluster extent. If no significant clusters were observed at this threshold, an exploratory threshold of p<0.001 (uncorrected) was applied to identify potential effects. We acknowledge the relatively small sample size may have limited statistical power to detect whole-brain effects.

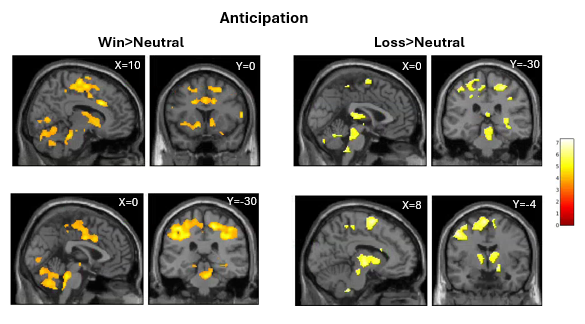
**Supplementary Figure 2. Brain activation patterns resulting from the exploratory whole brain analysis for the NU group in the anticipation phase.**

| **Condition** | **Contrast** | **Clusters** | **N of voxels** | **Peak coordinates** |
| --- | --- | --- | --- | --- |
| **Anticipation**  **Win vs. Neutral**  **Loss vs. Neutral**  **Feedback**  **Win Hit vs. Neutral**  **Loss Hit vs. Neutral** | CU>NU  NU>CU  CU>NU  NU>CU  CU>NU  NU>CU  CU>NU  NU>CU | /  Brainstem  Frontal Pole  /  Brainstem  /  Brainstem  / | /  k=221  k=131  /  k=157  /  k=151  / | /  x=0, y=-38, z=-38  x=16, y=42, z=42  /  x=10, y=-42, z=-30  /  x=-2, y=-34, z=-14  / |

**Supplementary Table 1: Significant group differences between CU and NU during anticipation and feedback phases. Results are reported for both directions of comparison (CU>NU; NU>CU), cluster size (k, number of voxels) and peak MNI coordinates for each cluster, with names of main areas covered.**
